# A Field-Side Triage Model for Early Specialist Referral After Acute Lower Extremity Sports Injuries in Young Athletes: Development and Internal Validation

**DOI:** 10.64898/2026.03.18.26348754

**Authors:** Shinsuke Sakoda, Hiroto Kumagae, Kimiaki Kawano

## Abstract

**Objective:** To develop and internally validate a field-side triage model to support early specialist referral decisions in young athletes with acute lower extremity sports injuries, where diagnostic resources are often limited.

**Design:** Retrospective cohort study.

**Setting:** Single-center sports medicine clinic.

**Participants:** Athletes aged ≤22 years presenting with acute lower extremity sports injuries between January 2017 and November 2025.

**Independent Variables:** Age, sex, functional severity, injury site, and injury mechanism assessed at initial presentation.

**Results:** A total of 2,129 athletes were included, with 276 (13.0%) undergoing surgery. Independent predictors were older age, female sex, greater functional severity, knee involvement, and high-energy deceleration mechanisms. The full model showed good performance (AUC 0.890; Brier score 0.073; calibration slope 1.00), and the simplified model also demonstrated high discrimination (AUC 0.883). Risk stratification showed increasing surgical rates across low-, intermediate-, and high-risk groups. Decision curve analysis demonstrated greater net benefit than treat-all and treat-none strategies across clinically relevant thresholds.

**Conclusions:** A field-side prediction model based on readily obtainable clinical variables demonstrated good performance for identifying young athletes at risk of requiring surgical intervention and may support early specialist referral decisions in resource-limited settings.

**Clinical Relevance:** This model provides a practical tool for early risk stratification using simple clinical information, supporting timely and appropriate referral decisions in field-side and initial clinical settings.

## Introduction

Timely specialist referral is a critical component of effective sports injury management. In young athletes, delayed recognition of serious injuries may lead to prolonged recovery, preventable complications, and delayed return to play. Conversely, excessive referral of minor injuries can overburden specialist services and increase healthcare costs. Achieving an appropriate balance between timely identification of clinically important injuries and avoidance of unnecessary referrals remains a key challenge in sports medicine. ^1–4^

A dynamic model of injury etiology has highlighted the complex interaction between intrinsic and extrinsic factors in sports injuries. ^1–3^ In many real-world sports settings, initial injury assessment occurs in environments where diagnostic resources are limited. Field-side evaluations are often performed by athletic trainers, coaches, or team medical staff using only a small amount of clinical information immediately after injury. ^5–7^ In such situations, referral decisions frequently rely on subjective judgment and varying levels of clinical experience, which may result in inconsistent triage practices. The development of simple and objective approaches to guide early referral decisions could therefore improve both the efficiency and consistency of injury management.

Several clinical decision rules have been developed to support early management of musculoskeletal injuries. For example, the Ottawa ankle and knee rules were designed to guide radiographic imaging decisions following acute injury and have substantially improved diagnostic efficiency in emergency medicine settings. ^8–10^ However, these tools were developed primarily to detect fractures and guide imaging utilization rather than to support early referral decisions across a broad range of sports-related injuries. In sports medicine practice, such referral decisions typically involve consultation with orthopedic surgeons or sports medicine specialists for further diagnostic evaluation and management.

Clinical prediction models have increasingly been used to support medical decision-making by integrating multiple clinical variables into structured risk estimation. ^11–16^ Such models have been widely applied in diverse healthcare settings to assist with triage decisions and risk stratification. Nevertheless, few prediction models have been developed specifically to support early specialist referral in young athletes presenting with acute sports injuries using information that is readily obtainable during the initial stages of injury assessment.

Therefore, the purpose of this study was to develop and internally validate a practical risk stratification model designed to support early specialist referral decisions in young athletes presenting with acute lower-extremity sports injuries using clinical information available at the time of initial evaluation.

## Methods

### Study Design and Participants

This retrospective cohort study was conducted at a specialized sports medicine center. Young athletes aged ≤22 years who presented with acute sports-related injuries involving the thigh, knee, lower leg, ankle, or foot between January 2017 and November 2025 were eligible for inclusion. The study was conducted in accordance with the Transparent Reporting of a Multivariable Prediction Model for Individual Prognosis or Diagnosis (TRIPOD) statement. ^11, 12^ The study flow, model development, and internal validation process are summarized in Figure 1.

**Figure 1.**
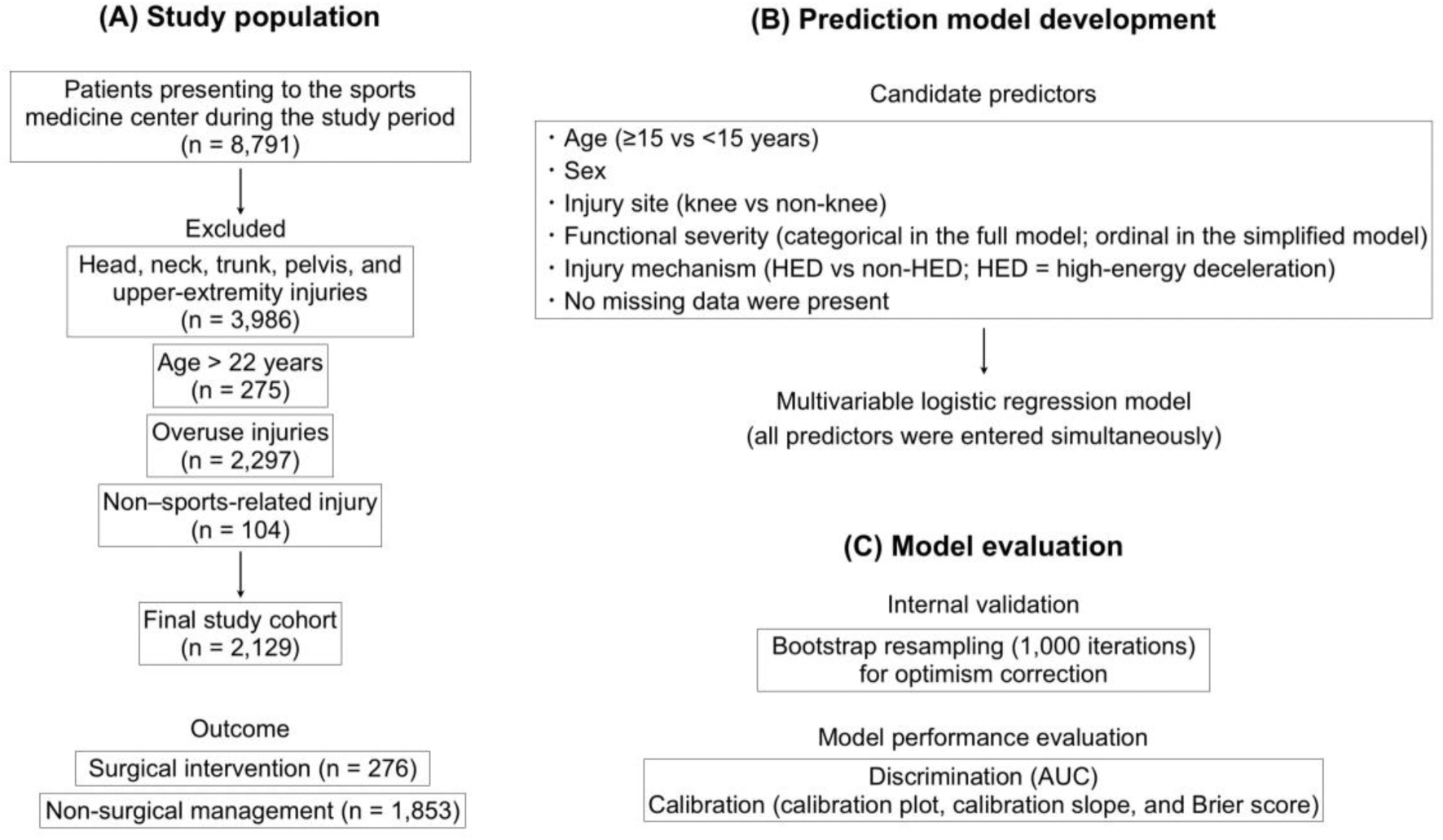
Study flow, model development, and internal validation of the referral-oriented triage model for acute lower-extremity sports injuries. (A) Study population. Patients presenting to the sports medicine center during the study period were screened. After excluding head, neck, trunk, pelvis, and upper-extremity injuries, patients aged >22 years, overuse injuries, and non–sports-related injuries, a final cohort of 2,129 athletes with acute lower-extremity injuries was included. The outcome was surgical intervention. (B) Prediction model development. Candidate predictors included age (≥15 vs <15 years), sex, injury site (knee vs non-knee), functional severity, and injury mechanism (HED vs non-HED). In the full model, functional severity was entered categorically with Grade 1 as the reference category; in the simplified model, it was entered as an ordinal variable. No missing data were present. (C) Model evaluation. Internal validation was performed using bootstrap resampling with 1,000 iterations to estimate optimism-corrected performance. Model performance was assessed in terms of discrimination using the area under the receiver operating characteristic curve (AUC) and calibration using calibration plots, calibration slope, and the Brier score.

### Outcome Measure

Surgical intervention was selected as the primary outcome because it represents a clinically meaningful and objective indicator of injury severity that can be consistently identified in retrospective datasets. Injuries requiring surgical management almost invariably necessitate specialist evaluation, making this outcome a pragmatic surrogate for conditions that require early orthopedic referral.

### Candidate Variables

Candidate predictors were selected based on clinical relevance and availability during early post-injury assessment, consistent with recommendations for clinical prediction model development. ^13–16^ The candidate variables comprised age, sex, injury site, functional severity, and injury mechanisms involving substantial weight-bearing load. To enhance real-world applicability, age was dichotomized at 15 years. Injury site was categorized as knee versus non-knee injuries.

### Functional Severity

Functional severity was classified into three grades according to activity limitation immediately after injury. Grade 1 was defined as the ability to continue sports participation immediately after the injury. Grade 2 was defined as inability to continue sports participation due to pain or functional limitation but able to walk independently. Grade 3 was defined as inability to bear weight on the injured limb and inability to walk independently immediately after the injury.

This classification was designed to capture observable functional impairment during early field-side or initial clinical assessment without requiring detailed physical examination or imaging.

### Injury Mechanism

Injury mechanisms were categorized according to whether substantial weight-bearing load was transmitted through the lower extremity at the time of injury.

Injuries occurring during high-energy deceleration movements involving substantial lower-extremity load transmission (e.g., landing, cutting, or rapid deceleration) were classified as high-energy deceleration (HED) mechanisms, irrespective of contact. All other injury mechanisms were categorized as non–high-energy deceleration (non-HED) mechanisms.

### Model Development

Model development followed a clinically oriented risk assessment framework designed to support early triage decisions in field-side settings.

The full model incorporated all candidate predictors available during early injury assessment, including age group (≥15 vs <15 years), sex, injury site (knee vs non-knee), functional severity, and injury mechanism (HED vs non-HED). In the full model, functional severity was included as a categorical variable with Grade 1 as the reference category to allow flexible estimation of effects across severity levels.

Because detailed information regarding the precise injury mechanism may not always be reliably obtained during early field-side or initial clinical assessment, a simplified model that excluded injury mechanism was additionally developed. In the simplified model, functional severity was treated as an ordinal variable (Grade 1–3) to reduce model complexity and enhance clinical applicability, assuming a monotonic increase in risk across severity levels.

Univariable logistic regression was first performed to evaluate associations between candidate variables and surgical intervention. Multivariable logistic regression was then used to construct the prediction model according to established approaches for clinical prediction model development. ^13–16^

To enhance clinical applicability in field-side settings, model performance using only readily obtainable variables (simplified model) was compared with that of the extended model incorporating injury mechanism.

### Model Performance and Clinical Utility

Model discrimination was assessed using the area under the receiver operating characteristic curve (AUC). Calibration was evaluated using calibration plots, the Brier score, and the calibration slope, which was estimated by fitting a logistic regression model with the observed outcome as the dependent variable and the linear predictor from the original model as the independent variable.

To assess internal model stability and potential overfitting, internal validation was performed using bootstrap resampling with 1000 iterations. Bootstrap-corrected estimates of model performance were obtained. ^13–16^

Clinical utility was assessed using decision curve analysis (DCA) to compare the net benefit of the model with treat-all and treat-none strategies across a range of threshold probabilities. ^17, 18^

Predicted probabilities were categorized into three referral-oriented risk groups using predefined thresholds of 5% and 15%. These thresholds were selected to reflect clinically meaningful referral decision points, balancing the need to minimize missed injuries requiring specialist management while avoiding excessive specialist referrals.

### Ethical Considerations

This study was approved by the institutional review board, and the requirement for informed consent was waived due to the retrospective nature of the study.

## Results

### Patient Characteristics

A total of 2,129 young athletes were included in the analysis. The overall surgical incidence was 13.0%. Athletes aged ≥15 years and female athletes demonstrated higher surgical rates. Knee injuries and greater functional severity based on post-injury activity limitation were strongly associated with surgical intervention (Table 1).

**Table 1.** Baseline characteristics of the study population.

| Variables | Cases (n) | Surgery (n) | Surgery (%) |
| --- | --- | --- | --- |
| Total | 2129 | 276 | 13.0 |
| Age group |  |  |  |
| < 15 years | 811 | 48 | 5.9 |
| ≥ 15 years | 1318 | 228 | 17.3 |
| Sex |  |  |  |
| Female | 533 | 103 | 19.3 |
| Male | 1596 | 173 | 10.8 |
| Injury site |  |  |  |
| Knee | 699 | 197 | 28.2 |
| Non-knee | 1430 | 79 | 5.5 |
| Severity |  |  |  |
| Grade 1 | 644 | 14 | 2.2 |
| Grade 2 | 1133 | 98 | 8.6 |
| Grade 3 | 352 | 164 | 46.6 |
| Injury mechanism |  |  |  |
| HED | 891 | 186 | 20.9 |
| Non-HED | 1238 | 90 | 7.3 |

Severity: Functional injury severity graded at initial assessment (Grade 1: able to continue sports activity; Grade 2: unable to continue sports but able to walk; Grade 3: unable to bear weight and unable to walk independently).
HED: High-energy deceleration mechanisms (e.g., landing, cutting, rapid deceleration).

### Univariable and Multivariable Analyses

Univariable analysis demonstrated significant associations between surgical intervention and all candidate variables.

In multivariable analysis, older age, female sex, knee injury, greater functional severity, and HED mechanism remained independent predictors of surgical intervention (Table 2).

**Table 2.** Univariable and multivariable logistic regression analyses for predictors of surgical intervention.

| Variable | OR | 95% CI | *p* value | Adjusted OR | 95% CI | *p* value |
| --- | --- | --- | --- | --- | --- | --- |
| Age ≥ 15 years | 3.33 | 2.40–4.60 | < 0.001 | 2.55 | 1.74–3.74 | < 0.001 |
| Female sex | 1.97 | 1.51–2.57 | < 0.001 | 1.58 | 1.12–2.23 | 0.009 |
| Knee involvement | 6.70 | 5.06–8.86 | < 0.001 | 7.64 | 5.47–10.66 | < 0.001 |
| Severity (vs Grade1) |  |  |  |  |  |  |
| Grade 2 | 4.26 | 2.41–7.50 | < 0.001 | 4.41 | 2.45–7.91 | < 0.001 |
| Grade 3 | 39.6 | 22.3–69.6 | < 0.001 | 34.49 | 18.82–63.19 | < 0.001 |
| HED | 3.37 | 2.57–4.41 | < 0.001 | 2.29 | 1.63–3.22 | < 0.001 |

Severity: Functional injury severity graded at initial assessment (Grade 1: able to continue sports activity; Grade 2: unable to continue sports but able to walk; Grade 3: unable to bear weight and unable to walk independently).
HED: High-energy deceleration mechanisms (e.g., landing, cutting, rapid deceleration).

Functional severity emerged as the strongest predictor, with markedly increased surgical risk observed in athletes with greater activity limitation immediately after injury.

### Model Performance

The bootstrap-corrected AUC was 0.890 for the full model and 0.883 for the simplified model, with corresponding Brier scores of 0.073 and 0.075. The simplified model also demonstrated high discrimination. The referral-oriented triage framework derived from the simplified model is shown in Figure 2.

**Figure 2.**
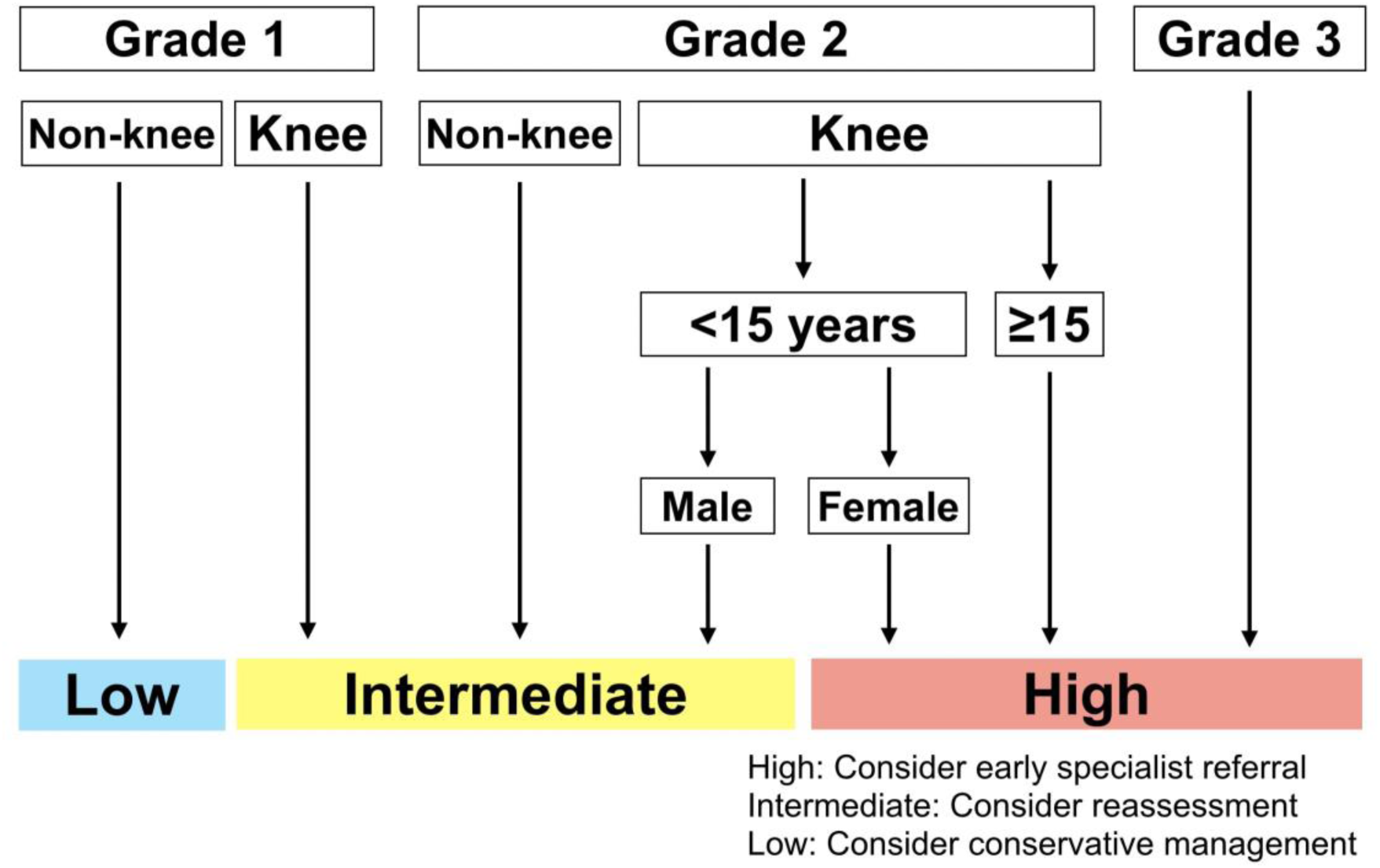
Referral-oriented triage framework derived from the simplified prediction model. The framework illustrates risk stratification based on predicted probabilities estimated from the multivariable logistic regression model using age, sex, knee involvement, and functional severity. Athletes are categorized into low-, intermediate-, and high-risk groups according to predefined probability thresholds (5% and 15%), corresponding to increasing likelihood of requiring surgical intervention.

Calibration assessment demonstrated good agreement between predicted and observed risks. The bootstrap-corrected calibration slope was 1.00, indicating good calibration without evidence of overfitting.

In the simplified model, functional severity was modeled as an ordinal variable (Grade 1–3). The regression coefficients are presented in Table 3, together with the corresponding logistic regression equation.

**Table 3.** Final simplified prediction model for surgical intervention.

| Predictor | Coding | $\beta$ coefficient | OR | 95% CI | *p* value |
| --- | --- | --- | --- | --- | --- |
| Age ≥ 15 years | 1 = age ≥ 15 years,<br>0 = age < 15 years | 0.981 | 2.67 | 1.83–3.89 | < 0.001 |
| Female sex | 1 = female,<br>0 = male | 0.659 | 1.93 | 1.39–2.69 | < 0.001 |
| Knee involvement | 1 = knee injury,<br>0 = non-knee | 1.936 | 6.93 | 5.02–9.57 | < 0.001 |
| Severity | Ordinal (1–3) | 2.063 | 7.87 | 5.99–10.33 | < 0.001 |
| Intercept | – | -8.226 | – | – | – |

The simplified model can be expressed as: $\text{logit}(p) = -8.226 + 0.981 \times (\text{Age} \geq 15 \text{ years}) + 0.659 \times \text{Female sex} + 1.936 \times \text{Knee involvement} + 2.063 \times \text{Severity}$ .

### Risk Stratification

Risk stratification categorized 1,150 athletes as low risk (surgical rate 1.7%), 393 as intermediate risk (7.6%), and 586 as high risk (38.6%) (Figure 3A). Using the high-risk category as a referral threshold yielded a sensitivity of 81.9%, a specificity of 80.6%, a positive predictive value of 38.6%, and a negative predictive value of 96.8%. At this threshold, the positive predictive value corresponds to a number needed to refer of approximately 2.6, indicating that roughly three referrals would identify one athlete requiring surgical intervention.

**Figure 3.**
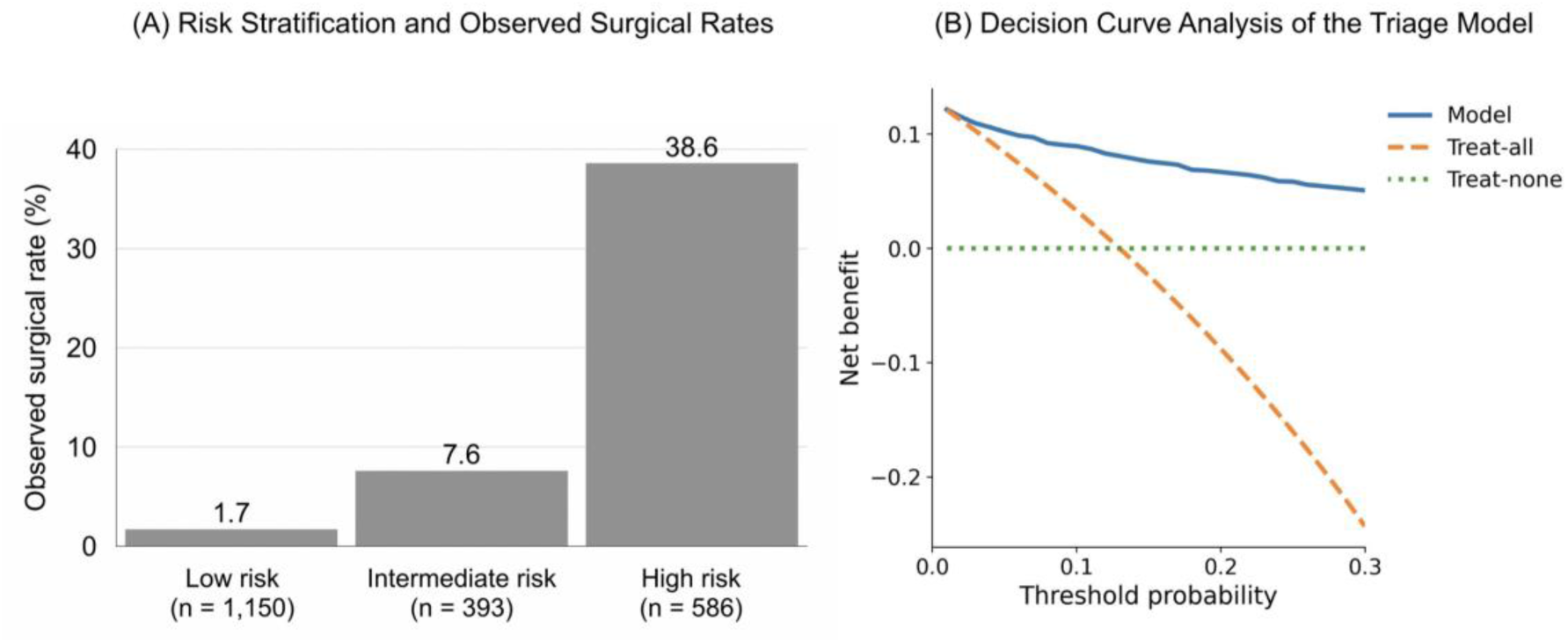
Risk Stratification and Clinical Utility of the Triage Model (A) Distribution of predicted probabilities and observed surgical rates across three risk categories (<5%, 5–15%, >15%). (B) Decision curve analysis demonstrating the net benefit of the triage model compared with treat-all and treat-none strategies across threshold probabilities.

Decision curve analysis (DCA) demonstrated greater net benefit than both treat-all and treat-none strategies across threshold probabilities of approximately 5% to 20% (Figure 3B).

## Discussion

A key contribution of this study is the introduction of a referral-oriented triage framework for acute sports-related injuries. ^19, 20^ Unlike existing clinical decision rules, such as the Ottawa ankle and knee rules, which were developed to guide imaging decisions primarily for fracture detection, the present model addresses a different clinical objective: early identification of athletes who may require specialist evaluation due to injuries with a high likelihood of surgical management. ^8, 9^ In real-world settings, including field-side assessments and primary care environments, referral decisions are often required before definitive diagnoses are established. By extending the concept of clinical decision rules from diagnostic or imaging triage to referral triage, the proposed model provides a structured approach to early identification of clinically significant injuries. ^19, 20^

In this study, we developed and internally validated a triage model for young athletes with acute lower-extremity sports injuries using information available at initial assessment. The model demonstrated good discrimination and calibration and enabled clinically meaningful risk stratification. Importantly, a simplified model based solely on readily observable variables maintained high predictive performance, supporting its potential applicability in resource-limited or field-side settings.

Within the broader literature on clinical prediction models, most previously reported models in sports medicine have focused on predicting specific diagnoses, injury recurrence, or long-term outcomes after established injuries. Similarly, studies examining predictors of surgical treatment in sports-related injuries are typically diagnosis-specific or rely on imaging findings. ^13–16^ Although these approaches are valuable once diagnostic information becomes available, they are less applicable during the earliest stages of injury assessment when clinicians must make triage decisions based on limited information. In contrast, the present model intentionally relies on simple clinical observations obtainable immediately at presentation, allowing early risk stratification even before imaging or specialist evaluation, with surgical intervention serving as a pragmatic surrogate for clinically significant injuries requiring specialist referral.

Another important feature of the proposed framework is its staged risk assessment structure. Initial risk estimation can be performed using readily observable information, such as injury location and functional status, without requiring detailed knowledge of injury biomechanics or specialized diagnostic tools. When additional contextual information becomes available, incorporation of injury mechanism may further refine risk prediction. This stepwise structure balances practicality with predictive performance and may facilitate implementation across a wide range of clinical and field environments.

Among the predictors included in the model, immediate functional impairment following injury emerged as the strongest determinant of surgical risk, with risk increasing according to greater post-injury activity limitation. Knee involvement also emerged as a strong predictor, likely reflecting the relatively high prevalence of structural knee injuries requiring operative management in young athletes. Age and sex remained independent predictors after adjustment, suggesting that developmental and biological factors may influence both injury patterns and treatment requirements.

The proposed risk stratification framework may also inform practical decision-making in early clinical settings. Athletes classified as high risk may warrant prompt referral for specialist evaluation, given the substantially increased likelihood of requiring surgical management. Those in the intermediate-risk group may benefit from closer clinical follow-up and reassessment, whereas athletes in the low-risk group may be initially managed conservatively with appropriate monitoring. These recommendations are intended to support, rather than replace, clinical judgment, and should be interpreted within the context of individual patient presentation and available resources.

Although many of the predictors identified in this study may appear intuitive to experienced sports medicine clinicians, early injury assessment in real-world sports environments is frequently performed by non-specialist providers. ^6, 7^ In such settings, clinical observations may not always be interpreted consistently. By translating commonly recognized clinical indicators into a structured triage framework, the present model may help standardize referral decisions and make expert clinical reasoning more accessible in field-side environments.

Importantly, the simplified model demonstrated only slightly lower performance than the full model while maintaining high discrimination, indicating that accurate risk stratification may be achievable using only a small set of readily observable clinical variables. This finding enhances the potential applicability of the proposed triage framework in field-side environments where detailed information about injury mechanisms may not always be available.

Several limitations should be acknowledged. First, the study was conducted at a single center, which may limit the generalizability of the findings to other clinical settings or athletic populations. External validation in independent cohorts will therefore be necessary to determine the transportability of the proposed model. Second, the model was developed using retrospectively collected clinical data, and prospective validation will be required to evaluate its performance in real-time clinical practice. Third, the model was intentionally designed as a simplified triage tool based on readily obtainable clinical information. As a result, potentially relevant predictors such as imaging findings, detailed physical examination findings, or sport-specific biomechanical characteristics were not incorporated and may further improve predictive performance in more specialized settings. Finally, although decision curve analysis suggested potential clinical utility, the real-world impact of implementing this triage framework on referral patterns, healthcare utilization, and patient outcomes remains to be determined.

Despite these limitations, this study introduces a clinically practical and field-side framework for early referral triage in young athletes with sports injuries. By integrating easily obtainable clinical information into a structured risk stratification system, the proposed model may support more objective and reproducible referral decisions during early injury assessment. Future research should focus on external validation, prospective evaluation, and implementation studies to determine whether this field-side triage approach can improve clinical decision making and care pathways for injured athletes.

## Conclusion

A prediction model based on field-side clinical information demonstrated good discrimination and clinically meaningful risk stratification for identifying athletes at risk of requiring surgical intervention. By integrating a small set of readily observable clinical variables, the proposed triage framework may support early referral decisions for team physicians, athletic trainers, and other clinicians involved in field-side and early injury assessment.

## Data Availability

All data produced in the present study are available upon reasonable request to the authors

---

**Supplemental Table S1.** Detailed Injury Site and Mechanism.

| Category | Cases (n) | Surgery (n) | Surgery rate (%) |
| --- | --- | --- | --- |
| Injury site |  |  |  |
| Thigh | 238 | 2 | 0.8 |
| Knee | 699 | 197 | 28.2 |
| Lower leg | 107 | 7 | 6.5 |
| Ankle | 816 | 53 | 6.5 |
| Foot | 269 | 17 | 6.3 |
| Injury Mechanism |  |  |  |
| HED without contact | 607 | 135 | 22.2 |
| HED with contact | 284 | 51 | 18.0 |
| Non-HED without contact | 446 | 27 | 6.1 |
| Non-HED with contact | 792 | 63 | 8.0 |

HED: High-energy deceleration mechanisms (e.g., landing, cutting, rapid deceleration).
Contact indicates direct player-to-player or player-to-object contact at the time of injury.

